# Age-Related Changes in the Cholinergic System in Adults with Down Syndrome Assessed Using [^18^F]-Fluoroethoxybenzovesamicol Positron Emission Tomography Imaging

**DOI:** 10.1101/2024.10.28.24316136

**Authors:** Jason K Russell, Alexander C. Conley, Brian D. Boyd, J. Patrick Begnoche, Rachel Schlossberg, Allison Stranick, Adam J. Rosenberg, Lealani Mae Y Acosta, Dann Martin, Yasmeen Neal, Prabesh Kanel, Roger L. Albin, Michael S. Rafii, Julie Dumas, Paul A. Newhouse

## Abstract

Adults with Down syndrome are genetically predisposed to developing Alzheimer’s disease after the age of 40. The cholinergic system, which is critical for cognitive functioning, is known to decline in Alzheimer’s disease and although first investigated in individuals with Down syndrome 40 years ago, remains relatively understudied. Existing studies suggest individuals with Down syndrome have an intact cholinergic system at birth that declines through adulthood alongside the development of Alzheimer’s disease pathology. The present study provides the first description of cholinergic terminals in vivo in non-demented adults with Down syndrome utilizing [^18^F]-fluoroethoxybenzovesamicol PET imaging. In addition, we investigated age-associated decline in cholinergic terminal density. Sixteen non-demented adults with Down syndrome and 20 neurotypically developed individuals were studied, comparing radiotracer uptake groupwise and associations with age utilizing a voxel-based approach. Adults with Down syndrome displayed significantly increased [^18^F]-fluoroethoxybenzovesamicol uptake in the cerebellum, brainstem, thalamus, and numerous cortical regions compared to age-matched controls. Cholinergic terminal density in numerous cortical regions showed a steeper decline associated with increasing age in adults with Down syndrome than observed in neurotypically developed adults in the age range tested. These data suggest increased cholinergic terminal density in early adulthood in individuals with Down syndrome with a more rapid or earlier age-associated decline than is observed in neurotypically developed individuals.

## 1. Introduction

Alterations in the cholinergic system in individuals with Down syndrome (DS) have been investigated since the early 1980s (Brooksbank et al., 1989; Casanova et al., 1985; Kish et al., 1989). The cholinergic system is vital in modulating normal cognitive function (Graef et al., 2011), and decline in this system is responsible for impairments in cognitive functioning in aging and neurodegenerative disease (Dumas & Newhouse, 2011). Disruptions in cholinergic signaling have been hypothesized to play a role in the developmental disabilities observed in DS and the age- and Alzheimer’s disease (AD)-related decline in cognition observed in adults with DS (Casanova et al., 1985; Fodale et al., 2006). In autopsy studies in infants, it was found that the activity of the cholinergic enzyme acetylcholinesterase across multiple brain areas was similar between individuals with DS and non-DS controls (Brooksbank et al., 1989; Kish et al., 1989). Consistent with these data suggesting an intact cholinergic system, efforts to enhance cognition in children with DS using acetylcholinesterase inhibitors have been inconclusive, displaying modest or no effect on cognitive function (Kishnani et al., 2010; Spiridigliozzi et al., 2007, 2016).

As neurotypical individuals age, the cholinergic system declines (Albin et al., 2018; M. J. Grothe et al., 2014; Kanel et al., 2022), which is associated with decreased cognitive function (Richter et al., 2014). In neurotypical individuals, age-related cholinergic decline is accelerated by the development of AD pathology. Specifically, degeneration of the basal forebrain system (Ch1-4) has been demonstrated in AD (Dumas & Newhouse, 2011; Fernández-Cabello et al., 2020; Whitehouse et al., 1981, 1982), with more recent studies suggesting the decline of this system is present in subjective cognitive decline, a risk factor for the development of mild cognitive impairment (MCI) (Nemy et al., 2023; Scheef et al., 2019). Reduced cholinergic basal forebrain volume has been associated with poorer performance on numerous cognitive endpoints in healthy elderly individuals and individuals with AD (Richter et al., 2014; Xia et al., 2022).

Individuals with DS are virtually certain to develop AD (Fortea et al., 2021) and are reported to display a general accelerated aging phenomenon (Zigman, 2013). The triplicated chromosome 21 contains the gene for the amyloid precursor protein (APP), with overexpression of APP believed to underlie the early widespread amyloid deposition seen in individuals with DS (Rafii et al., 2019; Wisniewski et al., 1985). The life expectancy for individuals with DS has greatly increased over the last several decades and was reported as 57 years in 2019 (Iulita et al., 2022). As such, AD is now the leading cause of death in individuals with DS (Hithersay et al., 2019), with an estimated age of AD dementia onset of 53.8 years and an estimated age of death due to AD of 58.4 years (Iulita et al., 2022). Central cholinergic degeneration occurs early in the development of AD pathology and underlies many of the cognitive deficits observed in AD. In contrast to studies in children with DS, in adults with DS, some studies with acetylcholinesterase inhibitors have displayed modest positive effects on cognition (Heller et al., 2004; Spiridigliozzi et al., 2007), although effects are still mixed with no effects observed in young adults (Kishnani et al., 2009). Given the high risk of AD in individuals with DS, the role of basal forebrain cholinergic decline in AD symptomatology, and the accelerated aging phenomenon observed in numerous systems in AD, it is important to understand how the cholinergic system changes with aging in adults with DS.

Studies in adults with DS are relatively limited; one small study suggests a reduction in the number of neurons within the nucleus basalis of Meynert (Ch4) compared to controls, with an increased decline from 16-56 years of age in adults with DS (Casanova et al., 1985). More recent work in adults with DS, performed using magnetic resonance imaging (MRI) volumetry, indicated decreasing anteromedial (Ch1-3) and posterior (Ch4) basal forebrain volumes with the progression from asymptomatic through prodromal AD to symptomatic AD (Rozalem Aranha et al., 2023). These data indicate an important role for basal forebrain cholinergic degeneration alongside the development of AD pathology, which may underlie AD-related cognitive impairment in individuals with DS, as is observed in the typically developed population. Whether performed using MRI methodologies or post-mortem, cholinergic basal forebrain anatomy provides important information about the basal forebrain projection system in general; however, it does not indicate potential regional changes in cholinergic terminals. Assessments of changes in cholinergic terminals in adults with DS have not been performed. Such studies investigating regional cholinergic terminals would provide greater insight into cholinergic decline in adults with DS compared to neurotypical individuals with AD.

The density of the central cholinergic nerve terminals can be assessed using positron emission tomography (PET) imaging. Specifically, [^18^F]-fluoroethoxybenzovesamicol ([^18^F]-FEOBV) is a vesamicol-derived radiotracer that binds to the vesicular acetylcholine transporter (vAChT) found in cholinergic nerve terminals (Mulholland et al., 1993, 1998). [^18^F]-FEOBV uptake has shown decreased binding in patients with AD (Aghourian et al., 2017), with an association between cognitive performance and [^18^F]-FEOBV uptake observed (Xia et al., 2022). Furthermore, in neurotypical individuals, regional [^18^F]-FEOBV uptake has been observed to decline with increasing age (Albin et al., 2018; Kanel et al., 2022; Okkels et al., 2023). These data suggest that [^18^F]-FEOBV PET is sensitive to changes in cholinergic terminals seen with normal aging and in the early stages of AD.

The present study is the first to describe cholinergic terminal density in adults with DS using cholinergic PET imaging. As such, we first investigated whether individuals with DS display differences in [^18^F]-FEOBV uptake compared to an age-matched neurotypical control group. Secondly, we perform exploratory analyses to assess whether adults with DS display a more rapid age-related decline in cholinergic terminal density than neurotypical controls.

## 2. Methods

### 2.1. Participants

Sixteen individuals with DS not qualifying for a diagnosis of mild cognitive impairment or dementia, with an average age of 35.5 years old (range 19-50 years old, 8 male, 8 female), were recruited at Vanderbilt University Medical Center. These participants underwent neuropsychiatric assessment (Down Syndrome Mental State Exam, with KBIT-2 used to establish premorbid intellectual disability), an [^18^F]-FEOBV PET scan, and an MRI scan (see Table 1 for a summary of participant demographics and cognitive scores). Participants over 25 years old were recruited from the Trial Ready Cohort – Down Syndrome (TRC-DS), a cohort study where participants undergo multimodal imaging and cognitive assessments (NCT04165109). The younger participants, aged 18-24 years, were recruited into a study examining cholinergic terminal density in DS (NCT05231798). These participants completed the same MRI sequences and cognitive measures as those in the TRC-DS study and the [^18^F]-FEOBV PET scan. The Vanderbilt University Medical Center Institutional Review Board approved both the TRC-DS and the cholinergic substudy, and participants or legally authorized representatives gave written informed consent in accordance with the Declaration of Helsinki.

**Table 1.** Participant Demographics.

|  | **Down Syndrome Cohort** | **Neurotypical Cohort** |
| --- | --- | --- |
|  | **Total n = 16** | **Total n =20** |
| **Male/Female (% Female)** | 8/8 (50%) | 10/10 (50%) |
| **Age (range)** | 35.5 (19-50) | 35.5 (21-61) |
| **Race** |  | Not recorded |
| White | 15 (93.7%) |  |
| Black or African American | 0 (0%) |  |
| Mixed | 1 (6.3%) |  |
| **IQ: KBIT (range)** | 48.3 (40-66) | NA |
| **DSMSE (range)** | 70.4 (41-88.5) | NA |

The control group was selected to have the same average age and sex distribution. It consisted of 20 neurotypically developed adults with an average age of 35.5 years old (range 20-61, 10 males, 10 females). Participants under 40 were recruited at the University of Michigan, data which has been used in previous studies of changes in [^18^F]-FEOBV binding in aging (Albin et al., 2018; Kanel et al., 2022), while participants over 50 were recruited at Vanderbilt University Medical Center and the University of Vermont (NCT04129060) for a separate study on central cholinergic health following menopause where a subset of participants underwent [^18^F]-FEOBV PET imaging alongside additional study procedures including an MRI (see Table 1 for participant demographics). No participants from the control group had histories of psychiatric or neurological disease, and all participants had a normal neurological exam at the time of the study. The Institutional Review Board at University of Michigan approved the study containing control participants under 40 years of age, while the study containing control participants over 50 years old was approved by the University of Vermont Institutional Review Board, with the [^18^F]-FEOBV PET imaging substudy approved by the Vanderbilt University Medical Center Institutional Review Board. All participants gave written informed consent in accordance with the Declaration of Helsinki. All 13 neurotypical participants from the group under 40 years old were utilized, while seven neurotypical participants from the group over 50 years old were selected to generate a control group with a matched average age and a balanced sex distribution. Study data from the Vanderbilt University Medical Center was collected and managed using REDCap electronic data capture tools hosted at Vanderbilt University Medical Center (Harris et al., 2009, 2019).

### 2.2. MRI and [^18^F]-FEOBV PET scan acquisition

For adults with DS, MRI scans were performed on a research-dedicated Philips 3.0T Ingenia Elition X (Philips Medical Systems, Best, the Netherlands) at Vanderbilt University Medical Center. T1-weighted scans with TR = 6.7 ms, TE = 3.1 ms, and a spatial resolution of 1 x 1 x 1.2 mm^3^ (170 slices and 334-second duration) were utilized for PET image registration.

For the neurotypical individuals, MRI scans were performed on a research-dedicated Philips 3.0T Ingenia CX (Philips Medical Systems, Best, the Netherlands) at Vanderbilt University Medical Center or the University of Vermont, or a Philips 3.0T Achieva system (Philips Medical Systems, Best, the Netherlands) at the University of Michigan. T1-weighted scans with TR = 6.3 ms, TE = 2.9 ms, and a spatial resolution of 1 x 1 x 1 mm^3^ (225 slices and 338-second duration) from the scans at Vanderbilt University Medical Center and TR = 9.8 ms, TE = 4.6 ms and a spatial resolution 1 x 1 x 1 mm^3^ (136 slices) from the scans at Michigan University were utilized for PET image registration.

Participants with DS and control participants from Vanderbilt University Medical Center received 6.5mCi ± 10% [^18^F]-FEOBV via a slow I.V. bolus, while control participants from Michigan received 7.8 - 8.6mCi as previously described (Albin et al., 2018). Data acquisition began after a 3-hour uptake; subjects underwent a 30-minute scan consisting of six 300-second frames with a voxel size of 2 mm isotropic and a field of view (FOV) of 256 mm. PET scans were performed using a Philips Vereos digital PET/CT system at Vanderbilt University Medical Center and an ECAT Exact HR+ PET tomograph (Seimens Molecular Imaging) at University of Michigan. PET images from both sites were reconstructed using the OSEM-3D method with Gaussian smoothing and resampled to generate matching voxel sizes.

### 2.3. PET Image Processing

PET images were motion-corrected using MCFLIRT in FSL. PetSurfer (https://surfer.nmr.mgh.harvard.edu/fswiki/PetSurfer) (Greve et al., 2014, 2016) was used to coregister [^18^F]-FEOBV PET images with structural T1 weighted MRI images and partial volume correct PET images with the region-based voxel-wise (RBV) method (Thomas et al., 2011), with smoothing to a 4mm full width at half maximum (FWHM). The PET images were normalized to the eroded supraventricular white matter, similar to previous publications with this tracer (Albin et al., 2018) (Analysis without partial volume correction produces similar results and can be seen in Supplemental Figures 1-4). Coregistered PET and structural MRI images were imported to Advanced Neuroimaging Tools (Cullen & Avants, 2018) in Python (ANTsPy (v.0.5.3) and ANTsPyNet (v0.2.8), Python v3.12.4). Brains were extracted from structural T1-weighted MRIs using the brain extraction tool in ANTsPyNet and then transformed into MNI-152 space using symmetric normalization with ANTsPy. PET images were transformed into MNI-152 space using the *fwdtransforms* function in ANTsPy.

Study-specific masks were generated using nilearn (v0.10.4) in Python (v3.12.4) (Abraham et al., 2014). T1-weighted MRI images in MNI-152 space were used to calculate a whole brain and white matter masks. The white matter mask was then inverted to produce a whole brain mask, excluding white matter for each subject. These masks were then combined on a probabilistic level. If a voxel was within the masked area for >30% of participants, it was included in the final mask for the specific analysis.

### 2.4. Voxel-based analysis and statistics

All voxel-based analysis was performed using nilearn. Groupwise comparisons between adults with DS and neurotypical adults were performed using a whole-brain voxel-wise general linear model with group as the variant of interest and sex, as defined by the sex assigned at birth, and age as covariates. A threshold of p<0.001 with a minimum cluster size of 50 was used to determine regions displaying a significant difference between groups. To assess age-related change in [^18^F]-FEOBV uptake, a general linear model was used in a whole-brain voxel-wise approach with age as the variant of interest and sex as a covariate; as these endpoints were exploratory, a more permissive p-value of p<0.005 with a minimum cluster size of 50 was used to determine significant age-related change with both positive and negative correlations evaluated. To compare the rate of age-related decline between the adults with DS and neurotypically developed individuals, a general linear model assessing the interaction between group and age was performed with a p-value of p<0.005 and cluster size 50. A whole-brain assessment of age-related change in the two groups was performed, where the beta-value for the association between age and [^18^F]-FEOBV uptake masked to include only negative associations was displayed to assist in the visualization of this analysis. For this, no significance threshold was used such that the beta value could be observed in all voxels. Results are visualized using the xjView toolbox (https://www.alivelearn.net/xjview).

## 3. Results

### 3.1 [^18^F]-FEOBV uptake in adults with DS is greater across numerous regions than seen in neurotypically developed individuals

Adults with DS displayed an increase in [^18^F]-FEOBV uptake across numerous brain regions when compared to age-matched neurotypically developed control participants (Figure 1). The increased [^18^F]-FEOBV uptake in adults with DS is most apparent throughout the cerebellum and brainstem regions. Increased uptake is also observed through numerous cortical and subcortical areas, with increased uptake bilaterally in regions of the thalamus, occipital cortex, temporal cortex, frontal cortex, insular cortex, and parietal cortex.

**Figure 1.**
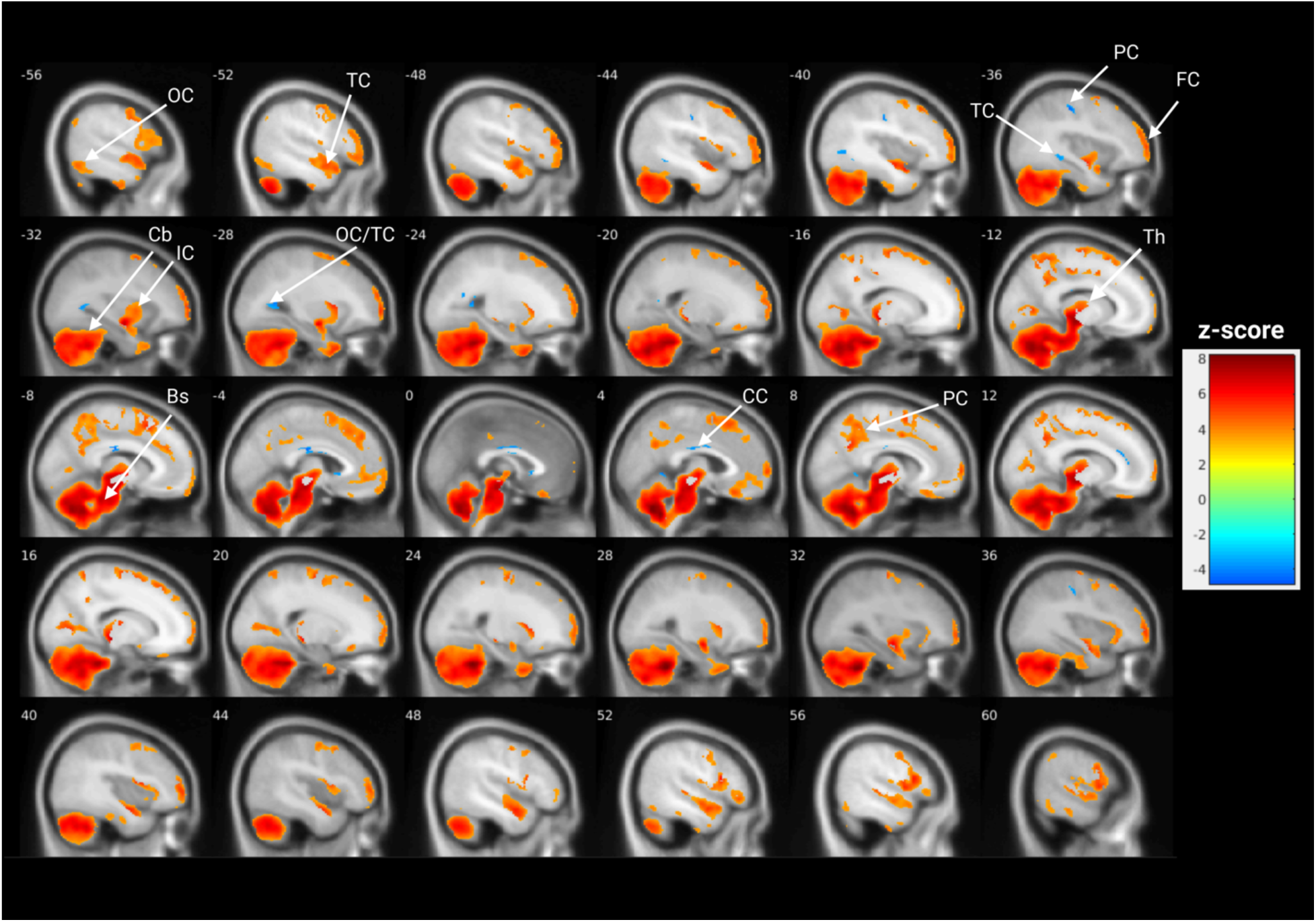
Adults with DS display increased [^18^F]-FEOBV uptake compared to age-matched neurotypically developed controls. Voxel-based comparison of [^18^F]-FEOBV uptake between adults with DS and neurotypical controls, adjusted for age and sex. Warm colors (green through red) indicate areas of significantly higher [^18^F]-FEOBV uptake in adults with DS, while cold colors (blues) indicate areas of significantly lower [^18^F]-FEOBV uptake in adults with DS. The color bar indicates the z-score. Bs, brainstem; Cb, cerebellum; CC, cingulate cortex; IC, insular cortex; FC, frontal cortex; OC, occipital cortex; PC, parietal cortex; TC, temporal cortex; Th, thalamus.

In addition to the diffuse regions of increased [^18^F]-FEOBV uptake in adults with DS, several restricted clusters display reduced [^18^F]-FEOBV uptake compared to neurotypically developed control individuals. Notably, clusters with decreased [^18^F]-FEOBV binding are observed in the cingulate cortex, the parietal cortex, and the boundary of the occipital and temporal cortices.

### 3.2 Adults with DS and neurotypically developed individuals display greater age-associated decreases in [^18^F]-FEOBV uptake than in neurotypically developed controls

Participants with DS display a number of clusters that have a negative association with increasing age, i.e., increasing age predicts lower [^18^F]-FEOBV uptake (Figure 2). These clusters are most predominant in cortical regions with numerous frontal, insular, parietal, cingulate, and temporal cortical clusters. A small cluster is also observed in the cerebellum. In addition to these negative associations, a small cluster in the frontal cortex displays an increase in [^18^F]-FEOBV uptake with increasing age.

**Figure 2.**
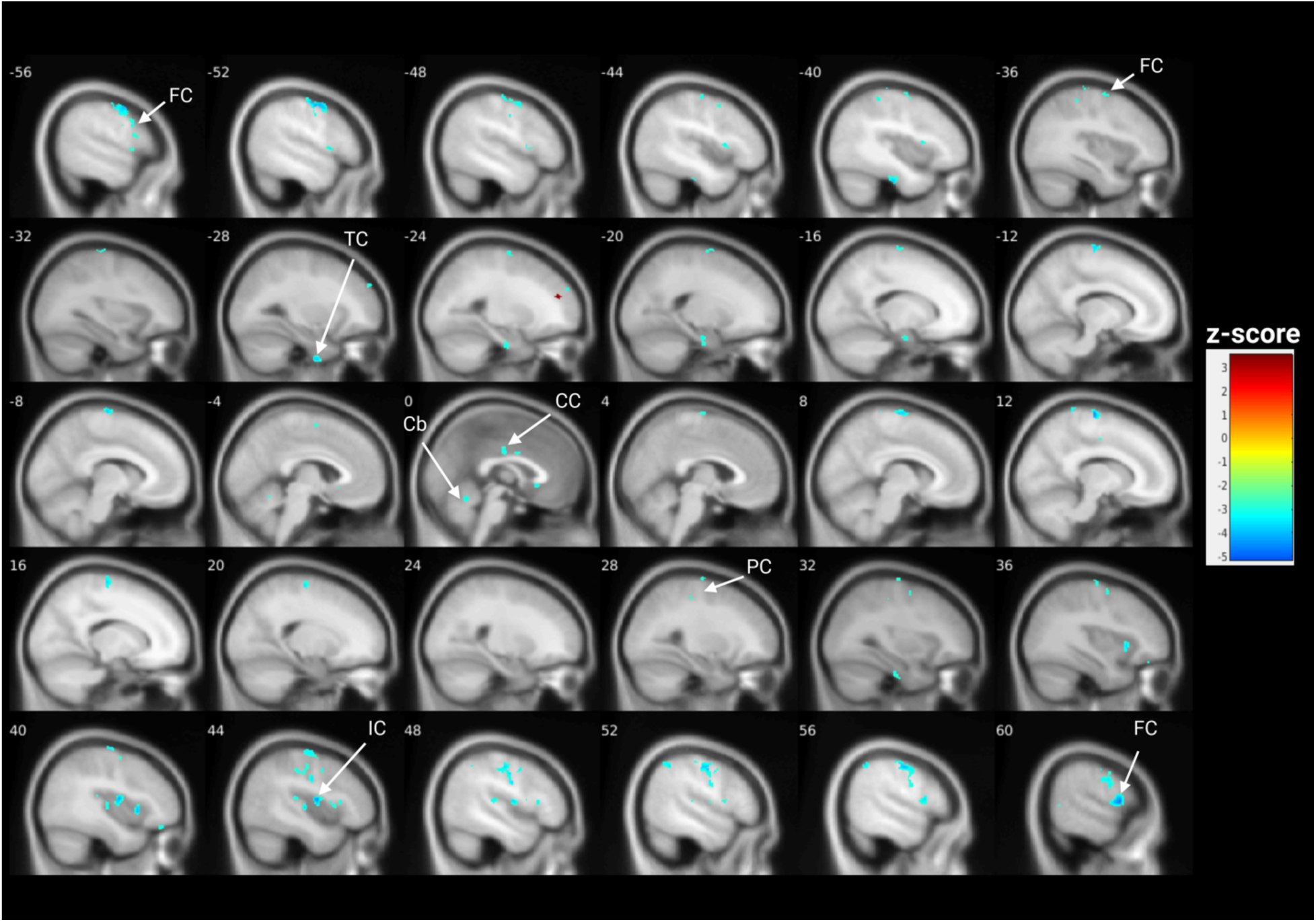
Adults with DS display age-associated decline in [^18^F]-FEOBV. Voxel-based association of [^18^F]-FEOBV uptake and age, adjusted for sex. Warmer colors (orange through red) indicate significantly increased [^18^F]-FEOBV binding with increased age, and colder colors (yellow through blue) indicate significantly decreased [^18^F]-FEOBV with increased age. The color bar indicates the z-score. Cb, cerebellum; CC, cingulate cortex; IC, insular cortex; FC, frontal cortex; PC, parietal cortex; TC, temporal cortex.

To assess whether adults with DS display a more rapid age-associated decline in cholinergic terminal density than age-matched neurotypically developed controls, the beta value for the relationship between age and [^18^F]-FEOBV uptake was evaluated across the whole brain for the two groups; neurotypically developed controls (Figure 3A) and adults with DS (Figure 3B). Subjectively, a range of areas display increased negative beta values in adults with DS. To quantify this, the age x group interaction was assessed (Figure 4). Adults with DS displayed a more negative age-associated decline in [^18^F]-FEOBV uptake than neurotypical controls in frontal, temporal, parietal, cingulate, insular, and occipital cortical regions. In addition to these cortical regions, increased age-associated decline was observed in cerebellar and brainstem clusters. One small cluster in the frontal cortex displayed a more negative age-related association in the neurotypically developed group.

**Figure 3.**
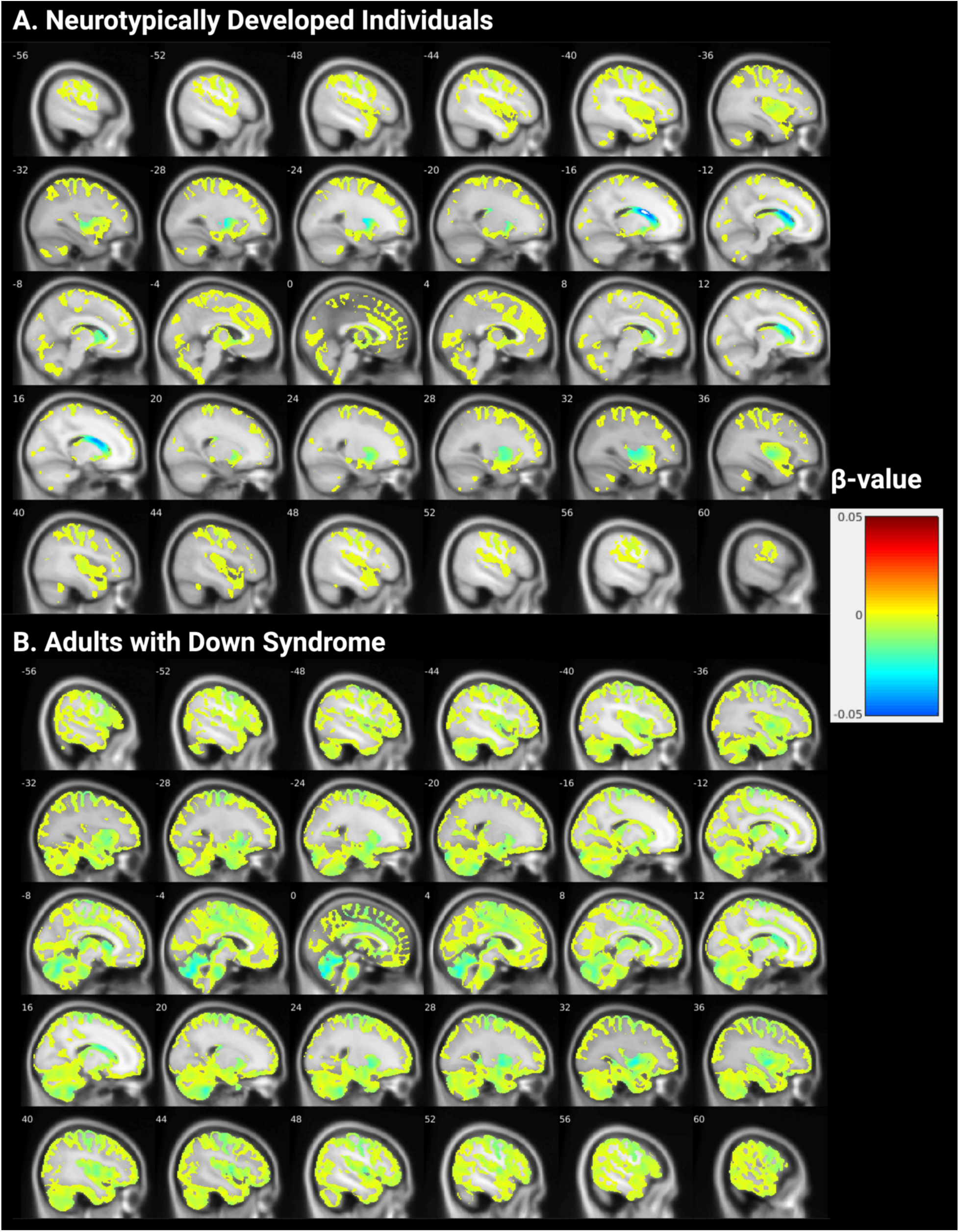
Age-associated changes in [^18^F]-FEOBV uptake. Shown are the negative beta-values of the whole brain (minus white matter) voxel-based association between age and [^18^F]-FEOBV uptake in neurotypically developed individuals (A) and adults with DS (B). Yellow indicates a beta value of zero, with colder colors indicating a decrease in [^18^F]-FEOBV uptake with increasing age. All negative voxels are shown with no masking by significance performed.

**Figure 4.**
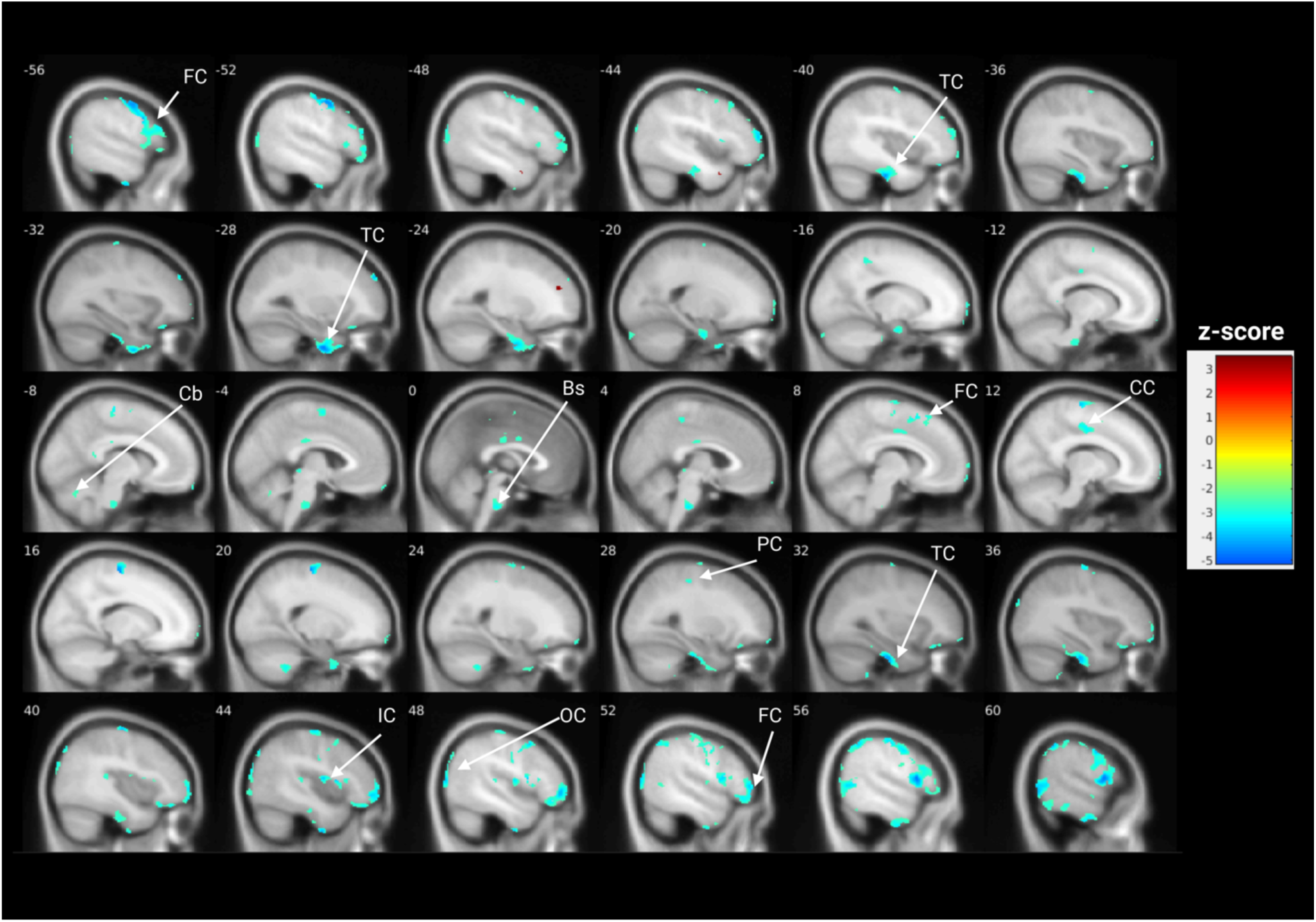
Adults with DS show increased age-associated decline in [^18^F]-FEOBV uptake compared to age-matched neurotypically developed controls. Voxel-based age x group interaction, colder colors (yellow through blue) indicate a significantly increased age-associated decline in adults with DS. In comparison, hotter colors (reds) indicate a significantly increased age-associated decline in the neurotypical control group. Bs, brainstem; Cb, cerebellum; CC, cingulate cortex; IC, insular cortex; FC, frontal cortex; OC, occipital cortex; PC, parietal cortex; TC, temporal cortex.

## 4. Discussion

The basal forebrain cholinergic system is known to decline early in the development of AD (Daamen et al., 2023; Scheef et al., 2019; Schmitz & Nathan Spreng, 2016), and loss of the cholinergic system is associated with the cognitive deficits observed in MCI and AD (Dumas & Newhouse, 2011; Richter et al., 2022; Xia et al., 2022). Studies in individuals with DS have suggested an intact cholinergic system at birth with a decline in the basal forebrain cholinergic system during adulthood as AD-related pathology develops. Prior work has focused on the basal forebrain nuclei either at post-mortem or using MRI volumetry (Casanova et al., 1985; Rozalem Aranha et al., 2023). The present study utilizes the [^18^F]-FEOBV PET ligand in adults with DS to assess the cholinergic terminal density in cortical and subcortical regions in relation to an age-matched control group and as a function of age. This approach allows for the assessment of projections from the basal forebrain, brainstem nuclei, and cholinergic interneurons. These assessments produce two predominant findings. Firstly, adults with DS have a higher level of [^18^F]-FEOBV uptake throughout large brain areas. Secondly, they display a more rapid age-associated decline in [^18^F]-FEOBV uptake than neurotypical control individuals. These contrasting findings suggest differing mechanisms affecting the cholinergic system in adults with DS, as discussed below.

Groupwise comparisons between two age-matched groups consisting of adults with DS and neurotypically developed adults revealed increased [^18^F]-FEOBV uptake, particularly in the cerebellum and brainstem, but also observed throughout numerous cortical and thalamic regions. Interestingly, the observed increase in [^18^F]-FEOBV uptake in a range of cortical regions in adults with DS is supported by MRI basal forebrain volumetry, where previous work has revealed increased posterior basal forebrain volume (which projects throughout cortical regions) in non-demented adults with DS compared to age-matched neurotypical controls (Rozalem Aranha et al., 2023). The reason for this increase in [^18^F]-FEOBV uptake in adults with DS compared to neurotypical controls is not immediately apparent. However, this may be a response to the early development of AD pathology that is observed in adults with DS (Rafii & Santoro, 2019). Previous studies have suggested an upregulation of cholinergic markers early in the development of AD (Dekosky et al., 2002). However, many observed changes are in subcortical regions not innervated by the cholinergic basal forebrain, the primary cholinergic system affected by AD pathology in neurotypically developed individuals. An alternative hypothesis is that this increased [^18^F]-FEOBV uptake may be an upregulation of cholinergic neurotransmission as compensation for developmental excitatory/inhibitory disturbances hypothesized in DS. *SLC18A3,* the gene encoding VAChT, is part of exon 1 of the choline acetyltransferase gene (*CHAT*) with evidence of coordinate regulation of their gene products (Gilmor et al., 1998). Increased regional VAChT density may thus indicate increased, compensatory cholinergic neurotransmission. Current hypotheses suggest an over-activation of the GABAergic system, resulting in excess inhibition early in development (Zorrilla de San Martin et al., 2018). One of the roles of the cholinergic system is to facilitate excitatory glutamatergic signaling in cortical (Baker et al., 2018; Marino et al., 1998) and cerebellar regions (Zhang et al., 2016); as such, the cholinergic system may display upregulation to compensate for this excess inhibition, resulting in a higher baseline level of cholinergic terminal density or signaling, as measured by [^18^F]-FEOBV uptake in adults with DS. Particularly notable increased regional cerebellar [^18^F]-FEOBV uptake was found broadly across cerebellar folia. Increased regional cerebellar [^18^F]-FEOBV uptake was seen also in early-moderate stage Parkinson disease subjects, perhaps as part of common compensatory cerebellar responses to forebrain pathologies (Brown et al., 2024; van der Zee et al., 2022). Previous work in non-demented adults with DS has reported increased glucose metabolic rate (GMR) measured by [^18^F]-Fluorodeoxyglucose ([^18^F]-FDG) PET during task-based conditions, in this case the increased GMR was predominant in the temporal/entorhinal cortex and hypothesized to be a compensatory mechanism for early AD pathology (Head et al., 2007).

As adults with DS get older, they appear to display age-associated decreases in [^18^F]-FEOBV uptake that is greater than is observed in neurotypically developed individuals across a similar age range. Regions displaying age-related decline within the frontal cortex and insular cortex overlap with clusters that displayed increased [^18^F]-FEOBV uptake in adults with DS when compared to neurotypically developed age-matched controls, while areas in the cingulate cortex overlapped with regions that displayed reduced [^18^F]-FEOBV uptake compared to age-matched controls. This suggests that a more rapid aging of the cholinergic system is occurring in adults with DS compared to neurotypical controls across the age range assessed while starting from a higher baseline [^18^F]-FEOBV uptake in some regions. Cholinergic terminal density has been described to decline with normal aging in the absence of neuropathology in neurotypically developed adults (Albin et al., 2018; M. Grothe et al., 2012; Kanel et al., 2022) with more rapid decline observed with the development of AD pathology (Aghourian et al., 2017; Nemy et al., 2023). Individuals with DS are considered to be on the AD continuum since birth (Jack et al., 2024), with evidence of AD pathology developing at age 12 years (Lemere et al., 1996). The early and rapid development of AD pathology in adults with DS (Fortea et al., 2021; Rafii & Santoro, 2019), with amyloid PET positivity by 40 years of age and a shortened interval from amyloid positivity to accumulation of tau tangles (Zammit et al., 2021, 2024), may underly this more rapid decline in cortical cholinergic terminal density in adults with DS compared to neurotypically developed controls.

This study provides evidence for a more rapidly declining cholinergic system from a higher baseline in adults with DS compared to neurotypically developed adults. However, it is not without limitations. Firstly, the sample size of 16 adults with DS is relatively small. However, this compares favorably to studies utilizing [^18^F]-FEOBV in studies of MCI (Xia et al., 2022), Parkinson’s disease (Horsager et al., 2022), and idiopathic REM sleep behavior disorder (Bedard et al., 2019). Somewhat larger studies have been completed with [^18^F]-FEOBV in neurotypically developed individuals (Albin et al., 2018; Kanel et al., 2022) and in Parkinson’s disease (Albin et al., 2022). Secondly, data from two sites was utilized in this study; the neurotypically developed control group data was predominantly collected at the University of Michigan, while the data from individuals with DS was collected at Vanderbilt University Medical Center. Reconstruction methods were matched to control for site differences, and images were resampled to match the larger voxel size from the data collected at the University of Michigan. Further corrections based on site were not performed due to demographic differences (sex and condition) in the sample between sites. Finally, individuals with Down syndrome are known to display neuroanatomical differences compared to neurotypically developed individuals (Beacher et al., 2010; Pujol et al., 2018). To control for these differences, we utilized study-specific probabilistic masks, and all analyses were performed following partial volume correction and warping to a common template.

Studies are ongoing to investigate the association between AD pathology and central cholinergic terminal density to differentiate whether the observed age-related reduction in [^18^F]-FEOBV uptake in the present cohort is related to an accelerated aging phenomenon in individuals with DS or the development of AD-related pathology. These studies will utilize data from the Alzheimer’s Biomarkers Consortium – Down Syndrome (ABC-DS) and TRC-DS studies. These studies include multimodal imaging assessments, including MRI, amyloid, and tau PET imaging. Amyloid and Tau PET imaging provide measures of Alzheimer’s pathology that can be associated with measures of cholinergic terminal density, including cholinergic basal forebrain volumetry from MRI imaging and [^18^F]-FEOBV PET. The relationship between cholinergic terminal density and different measures of Alzheimer’s disease pathology will be assessed at baseline and longitudinally.

## 5. Conclusion

We have presented data indicating that adults with DS have an increased baseline cholinergic terminal density with an increased age-associated decline compared to neurotypical controls. We demonstrated that adults with DS display increased [^18^F]-FEOBV uptake in cortical and sub-cortical regions compared to age-matched controls. In addition, we have presented data suggesting that the cholinergic system declines faster in adults with DS than in the neurotypical population. Ongoing studies will assess whether this more rapid cholinergic decline is associated with the early development of AD pathology. As this cohort increases in size, future studies will investigate the relationship between amyloid and tau pathology, cognitive function, and cholinergic decline as measured by [^18^F]-FEOBV in adults with DS. Data associating [^18^F]-FEOBV uptake with measures of AD pathology and cognitive endpoints may provide support for [^18^F]-FEOBV PET as a measure to aid in staging individuals with DS on the AD continuum and potential utility as an outcome measure in clinical trials.

## Data Availability

All data produced in the present study are available upon reasonable request to the authors

## Abbreviations

ABC-DS: Alzheimer’s Biomarker Consortium – Down Syndrome
AD: Alzheimer’s disease
ANTs: advanced normalization tools
APP: Amyloid precursor protein
Bs: brainstem
Cb: cerebellum
CC: cingulate cortex
Ch1-4: cholinergic nuclei 1-4
DS: Down syndrome
IC: insular cortex
FC: frontal cortex
[^18^F]-FEOBV: [^18^F]-fluoroethoxybenzovesamicol
FOV: Field of view
FSL: FMRIB Software Library
FWHM: full width at half maximum
KBIT-2: Kaufman Brief Intelligence Test Second Edition
MCI: Mild cognitive impairment
MRI: Magnetic resonance imaging
OC: occipital cortex
OSEM: Ordered subsets expectation-maximization
PC: parietal cortex
PET: Positron emission tomography
RBV: Region-based voxel-wise
TC: temporal cortex
TE: Echo time
Th: thalamus
TR: Repetition time
TRC-DS: Trial Ready Cohort – Down Syndrome
vAChT: vesicular acetylcholine transporter

## Acknowledgments

We would like to thank Alexandra Key and Elise McMillan from the Vanderbilt Kennedy Center for their assistance with recruitment and outreach, Sepideh Shokouhi for her input during the study’s design, and the participants and families for their continued dedication to science. The figures were assembled using BioRender.com.

## CRediT authorship contribution statement

**Jason K Russell:** Writing – original draft, Formal analysis, Conceptualization, Investigation, Visualization, Software, **Alexander C. Conley:** Writing – review and editing, Conceptualization, Funding acquisition, Investigation, **Brian D. Boyd:** Writing – review and editing, Formal analysis, Software, **J. Patrick Begnoche:** Writing – review and editing, Formal Analysis, Software, **Rachel Schlossberg:** Writing – review and editing, Investigation, Project administration, **Allison Stranick:** Writing – review and editing, Investigation, Project administration, **Adam J. Rosenberg:** Writing – review and editing, Resources, **Lealani Mae Y Acosta:** Writing – review and editing, Investigation, **Dann Martin:** Writing – review and editing, Investigation, **Yasmeen Neal:** Writing – review and editing, Investigation, **Prabesh Kanel:** Writing – review and editing, Resources, **Roger L. Albin:** Writing – review and editing, Resources, **Michael S. Rafii:** Writing – review and editing, Funding acquisition, **Julie Dumas:** Writing – review and editing, Funding acquisition, **Paul A. Newhouse:** Writing – review and editing, Funding acquisition, Supervision, Conceptualization

