## Supplemental Figures for "Age-Related Changes in the Cholinergic System in Adults with Down Syndrome Assessed Using [^18^F]-Fluoroethoxybenzovesamicol Positron Emission Tomography Imaging"

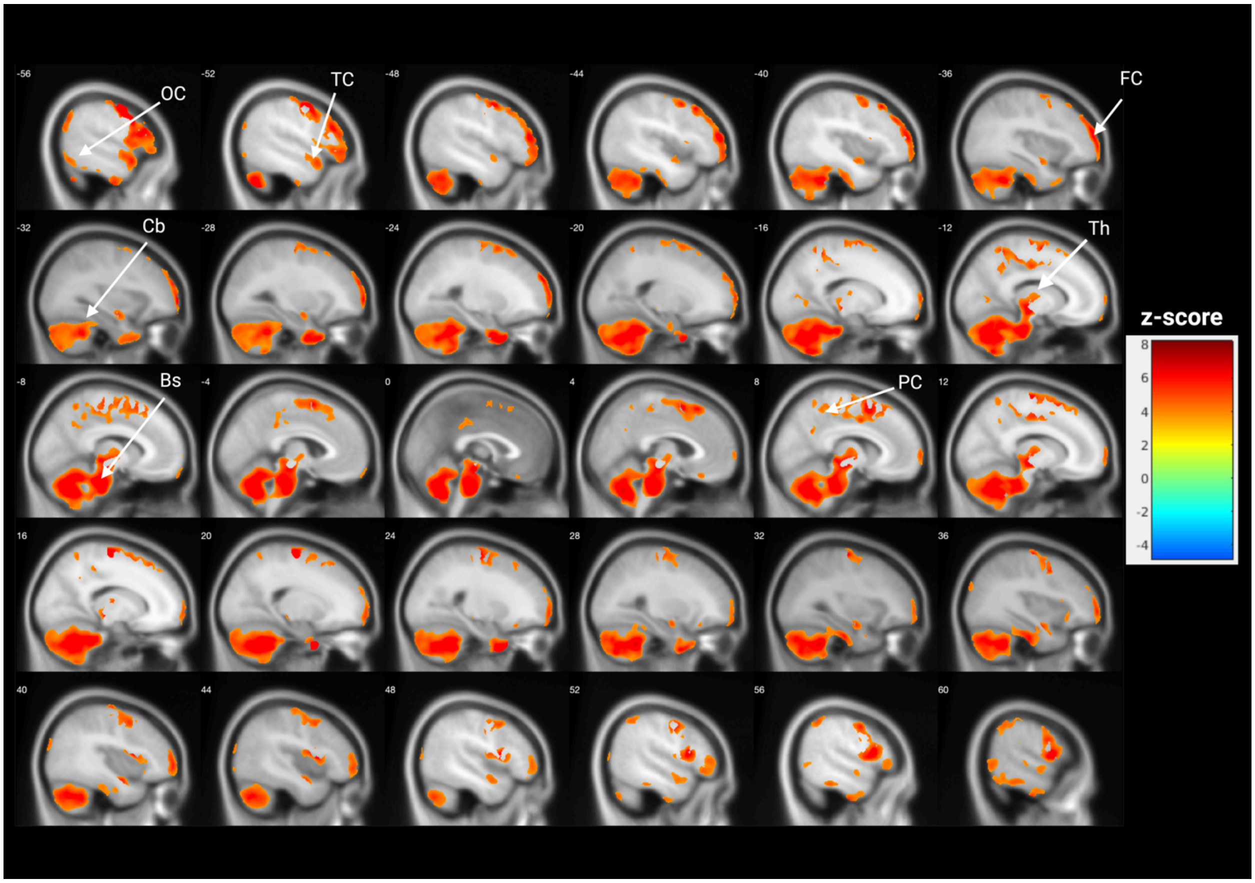


**Supplemental Figure 1. Adults with DS display increased [$^{18}$F]-FEOBV uptake compared to age-matched neurotypically developed controls when partial volume correction is not applied.** Voxel-based comparison of [$^{18}$F]-FEOBV uptake between adults with DS and neurotypical controls, adjusted for age and sex. Warm colors (green through red) indicate areas of significantly higher [$^{18}$F]-FEOBV uptake in adults with DS, while cold colors (blues) indicate areas of significantly lower [$^{18}$F]-FEOBV uptake in adults with DS. The color bar indicates the z-score. Bs, brainstem; Cb, cerebellum; FC, frontal cortex; OC, occipital cortex; PC, parietal cortex; TC, temporal cortex; Th, thalamus.

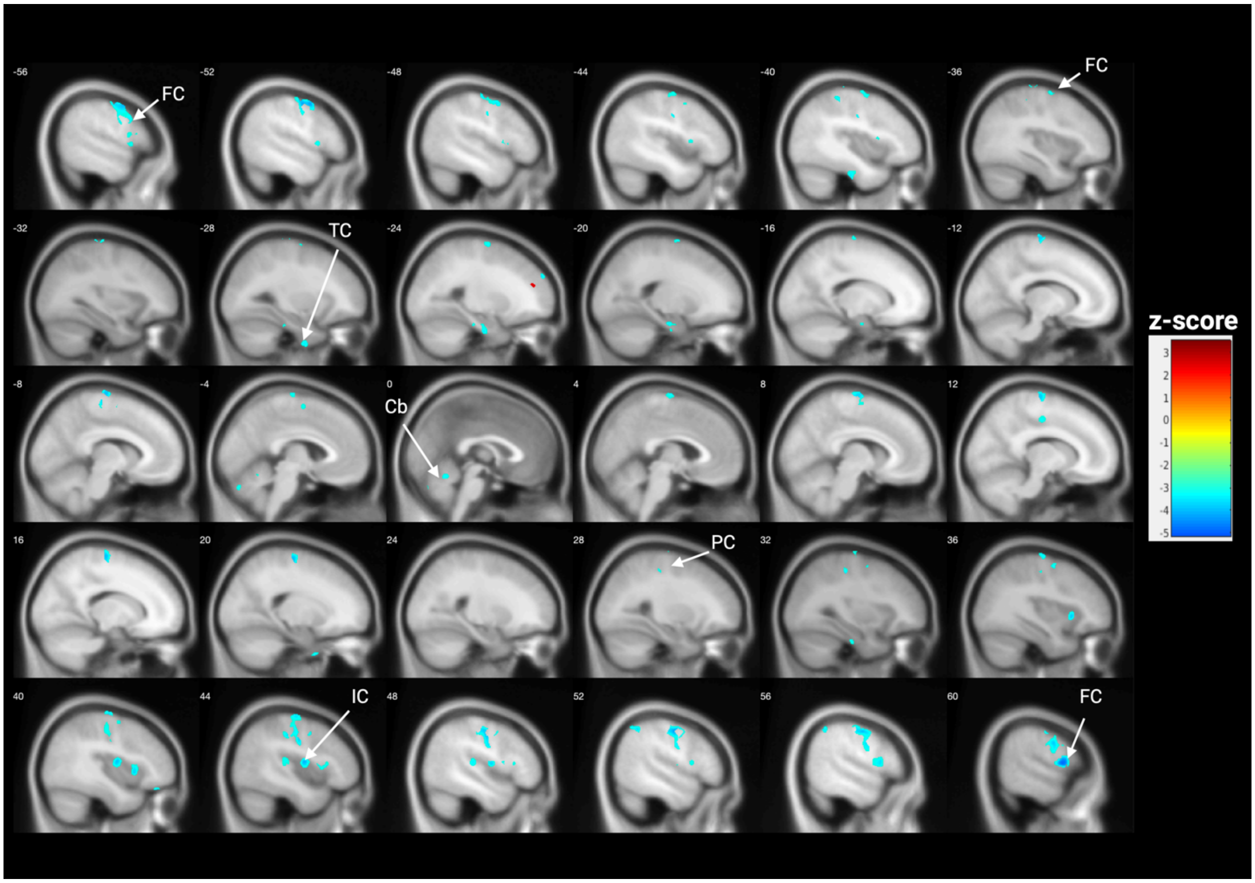


**Supplemental Figure 2. Adults with DS display age-associated decline in [$^{18}$F]-FEOBV with no partial volume correction applied.** Voxel-based association of [$^{18}$F]-FEOBV uptake and age, adjusted for sex. Warmer colors (orange through red) indicate significantly increased [$^{18}$F]-FEOBV binding with increased age, and colder colors (yellow through blue) indicate significantly decreased [$^{18}$F]-FEOBV with increased age. The color bar indicates the z-score. Cb, cerebellum; IC, insular cortex; FC, frontal cortex; PC, parietal cortex; TC, temporal cortex.

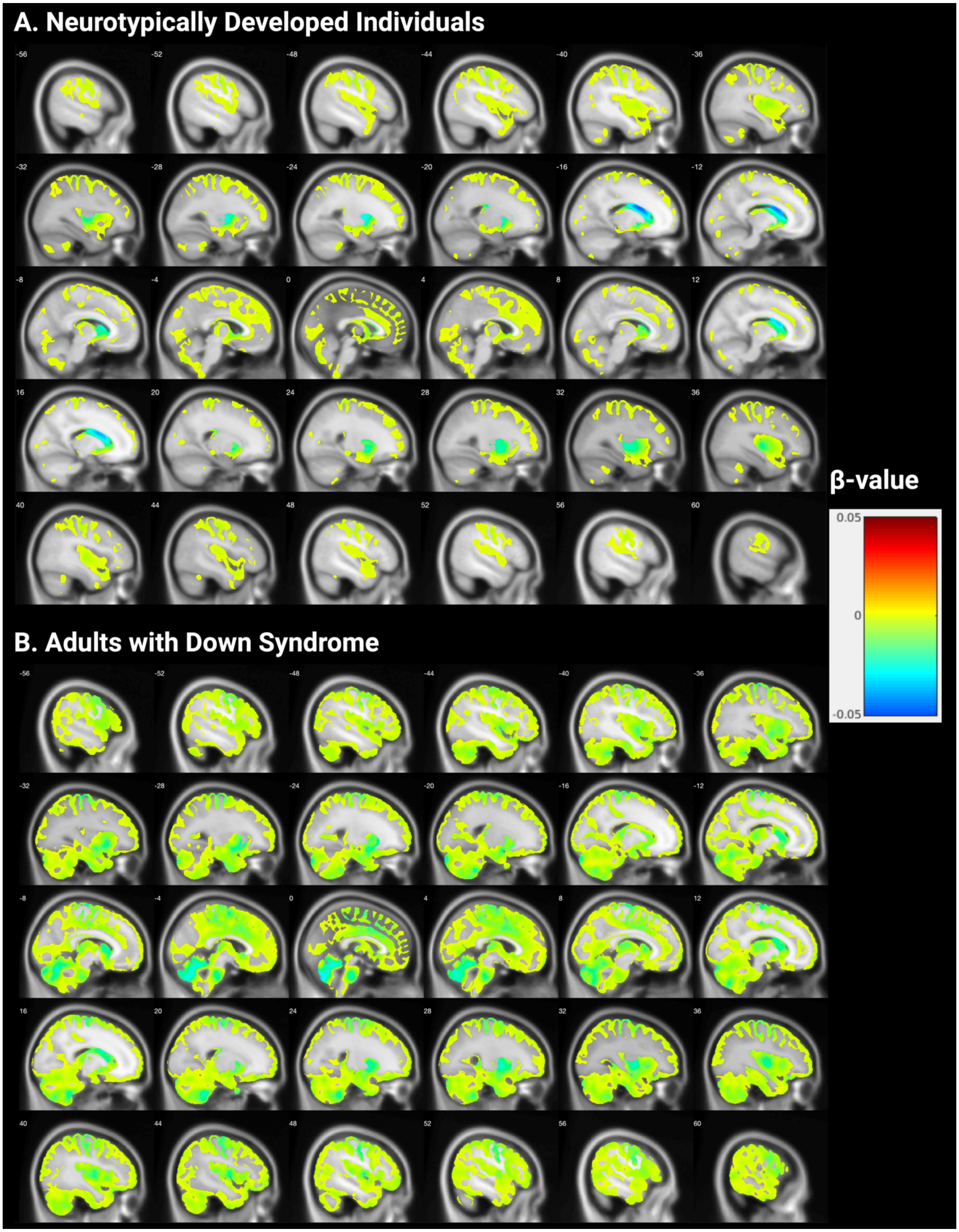


**Supplemental Figure 3. Age-associated changes in [$^{18}$F]-FEOBV uptake with no partial volume correction applied.** Shown are the negative beta-values of the whole brain (minus white matter) voxel-based association between age and [$^{18}$F]-FEOBV uptake in neurotypically developed individuals (A) and adults with DS (B). Yellow indicates a beta value of zero, with colder colors indicating a decrease in [$^{18}$F]-FEOBV uptake with increasing age. All negative voxels are shown with no masking by significance performed.

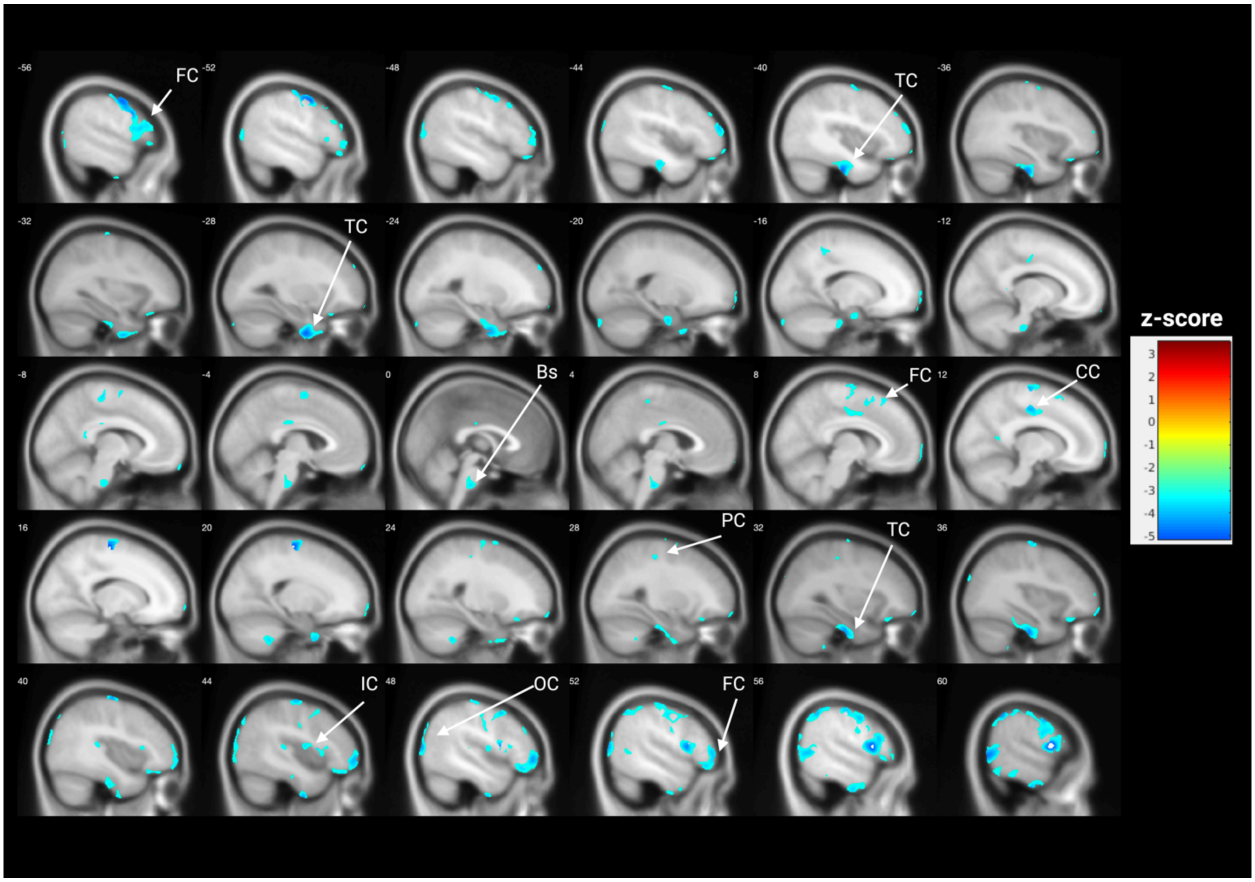


**Supplemental Figure 4. Adults with DS show increased age-associated decline in [$^{18}$F]-FEOBV uptake compared to age-matched neurotypically developed controls with no partial volume controlled.** Voxel-based age x group interaction, colder colors (yellow through blue) indicate a significantly increased age-associated decline in adults with DS. In comparison, hotter colors (reds) indicate a significantly increased age-associated decline in the neurotypical control group. Bs, brainstem; CC, cingulate cortex; IC, insular cortex; FC, frontal cortex; OC, occipital cortex; PC, parietal cortex; TC, temporal cortex.
